# Description of a community paediatric strategy offering a package of services to prevent malnutrition among children in one health district in Mali

**DOI:** 10.1101/2021.01.25.21250446

**Authors:** Thomas Roederer, Augusto E. Llosa, Susan Shepherd, Isabelle Defourny, Michel-Olivier Lacharité, Chibuzo Okonta, Mahamadou Magassa, Modibo Traoré, Myrto Schaefer, Rebecca F. Grais

**Affiliations:** Epicentre, 8 rue Saint Sabin, 75011, Paris France; Médecins Sans Frontières, Operational Center Paris, 75011, Paris France; ALIMA, Dakar, Sénégal; Ministry of Health, Bamako, Mali

## Abstract

**Background:** We present results from an intervention case study, the Soins Preventifs de l’Enfant (SPE) project, in Konséguéla health area, Mali. The intervention involved a network of community health workers providing a comprehensive preventive/therapeutic package, ultimately aiming at reducing under 24-month mortality. Associated costs were documented to assess the feasibility of replication and scale-up.

**Methods:** SPE program monitoring data were obtained from booklets specific to the program between 2010 and 2014. Data included sex, age, vaccination status, anthropometric measurements, Ready-To-Use-Supplementary Food distribution, morbidities reported by the mother between visits, hospitalizations over 18 months of follow-up. Cross-sectional surveys in the district of Koutiala, of which Konséguéla is one health area, were conducted yearly between 2010 and 2014 for comparison, using difference-in-difference approach. Ethical approval was granted from the Malian Ethical Committee.

**Results:** Global and Severe Acute Malnutrition prevalences decreased over time in Konséguéla as well as in the rest of the district, but the difference between areas was not significant. Children reaching 24 months were 20% less stunted in Konséguéla than children the same age outside (p<0.001). Mortality rates significantly decreased more in Konséguéla, while vaccination coverage for all antigens significantly increased in the meantime. The package cost approximately USD 95 per child per year; 56% of which was for the RUSF.

**Conclusion:** The results of this case study suggest a sustained impact of a community based, comprehensive health package on major child health indicators. Most notably, while improvements in acute malnutrition were found in the district as a whole, those in the intervention area were more pronounced. Trends for other indicators suggest additional benefits.

## Introduction

Global under 5 mortality decreased from 10 to 6.6 million deaths between 2000 and 2012[1,2]. Nonetheless, in 2012, sub-Saharan Africa accounted for 49% of the total deaths in children under the age of 5, compared to 33% in 2000. In Mali, under 5 mortality has decreased from 220 ‰ in 2000 to 176 ‰ in 2011[3]. However, reaching the MDG target of 85‰ by 2015, would have necessitated an unlikely recurring 50% yearly decrease in mortality[3]. National malnutrition figures in this age group have also failed to show significant progress, with 15% prevalence of wasting in 2000 and 13.4% in 2012[3,4,5]. The burden of malaria, pneumonia and diarrheal diseases, compounded by malnutrition, is still high, especially in the south of the country[5-10].

For over 10 years, efforts have been made to accelerate progress in child survival, vaccination coverage and growth indicators through provision of pediatric care packages. Thus far, the introduction of Integrated Management of Childhood Illness protocols in Malian health centers, associated with additional interventions such as bed-net and vitamin A distributions or breastfeeding promotion have not shown to decrease child mortality when compared to non-intervention districts[11-13]. Poor coverage and failure to address malnutrition specifically in the package are believed to explain the lack of benefit.

Interventions such as the distribution of ready to use supplementary foods (RUSFs), improving coverage of routine vaccination by using mobile teams as an extension of the Expanded Program on Immunization, and Seasonal Malaria Chemoprophylaxis (SMC) for prevention of malaria have shown meaningful progress in child health. Despite strong evidence to support each of these individual interventions, there is little published literature on the effects of combining these approaches on mortality reduction[14-29]. Here we present results from an intervention case study, the Soins Preventifs de l’Enfant (SPE) project, which took place in the seventeen villages comprising the Konséguéla health area CSCOM in Mali. The intervention involved community health workers networks and provided a comprehensive preventive/therapeutic package. This included reinforcement of the national EPI for all antigens within the schedule, distribution of Insecticide Treated bed Nets (ITN) and RUSF, in addition to early detection of malaria with treatment of simple cases and hospital referral of severe cases. Children within the program catchment area were also scheduled for 6 well-child visits between birth and 24 months of age. During these visits, anthropometric measurements, clinical evaluation and history of healthcare data were conducted and summarized in a health booklet, which was given to the mother of the child and used for better record-keeping.

The goal of this intervention was to reduce the under 24-month mortality rate by targeting several health indicators, including wasting, stunting, vaccination coverage for all EPI antigens, and reduction of malaria episodes. In parallel, associated costs were documented to assess the feasibility of replication and scale-up.

For comparability, annual cross-sectional surveys were conducted in the district, including the Konséguéla health area (SPE program). We report the first results of the SPE project after 4 years of implementation and compare health indicators among beneficiaries in Konséguéla and non-beneficiaries outside of Konséguéla.

## Methods

### Study Site

Médecins Sans Frontieres (MSF) is collaborating the Malian Ministry of Health (MoH) to provide a comprehensive package including capacity building, human resources, organization and logistical support to the local primary health structures (CSCOM by its French acronym) in Koutiala District, Sikasso Region, Mali.

The MSF interventions, including the SPE project, was implemented in Koutiala district, Sikasso region, southeastern Mali. This district is one of the most populated in Mali with approximately 580 000 inhabitants (of which 140 000 are under five years and 80 000 are under two). Recent surveys showed that wasting and stunting prevalences in under five in the Koutiala district were slightly above the national average (wasting: 16%, stunting: 44%), while malaria remains a major burden in under five children (33% with more than one episode in 2009)[9-10].

Konséguéla is one of the 42 health areas of the district, located in its western part, and is composed of 17 villages, with a population approx. 33 000 (8 000 under five years old and 3 000 under two).

### Interventions

In partnership with the MoH, the MSF program includes free pediatric consultations in the CSCOMS, following the national protocol. MSF is also reinforcing the EPI activities of the MoH at the CSCOMs level, by providing logistic support for vaccine storage and supply. To reinforce prevention, MSF started the SPE project in March 2010 in the Konséguéla health area, located in the western part of the Koutiala district. MSF outreach teams conducted monthly EPI reinforcement for all villages in the health area. During these visits, all newborns were identified and enrolled in the program. Mothers were also encouraged to enroll their infants in a growth-monitoring program based at the CSCOM, which is located in the city of Konséguéla, within 20 km of any of the 16 other catchment villages. Individual health booklets and mosquito nets were distributed to participating mothers and their child’s 6-month-of-age inclusion visit scheduled.

At the 6-month visit, the mother brought their child to the CSCOM, where his vaccination and nutritional statuses were assessed and the health booklet updated. The mothers then received a one-month ration of a complementary food supplement (Plumpy Doz, 250 kcal/day) for their child. Mothers were encouraged to return monthly for additional rations, and to bring the child every three months until they reach 12 months, then every 6 months between 18 and 24 months, the age of program discharge. All health care and nutritional supplements were provided free of charge. Figure 1 summarizes the timeline of the project.

**Figure 1.**
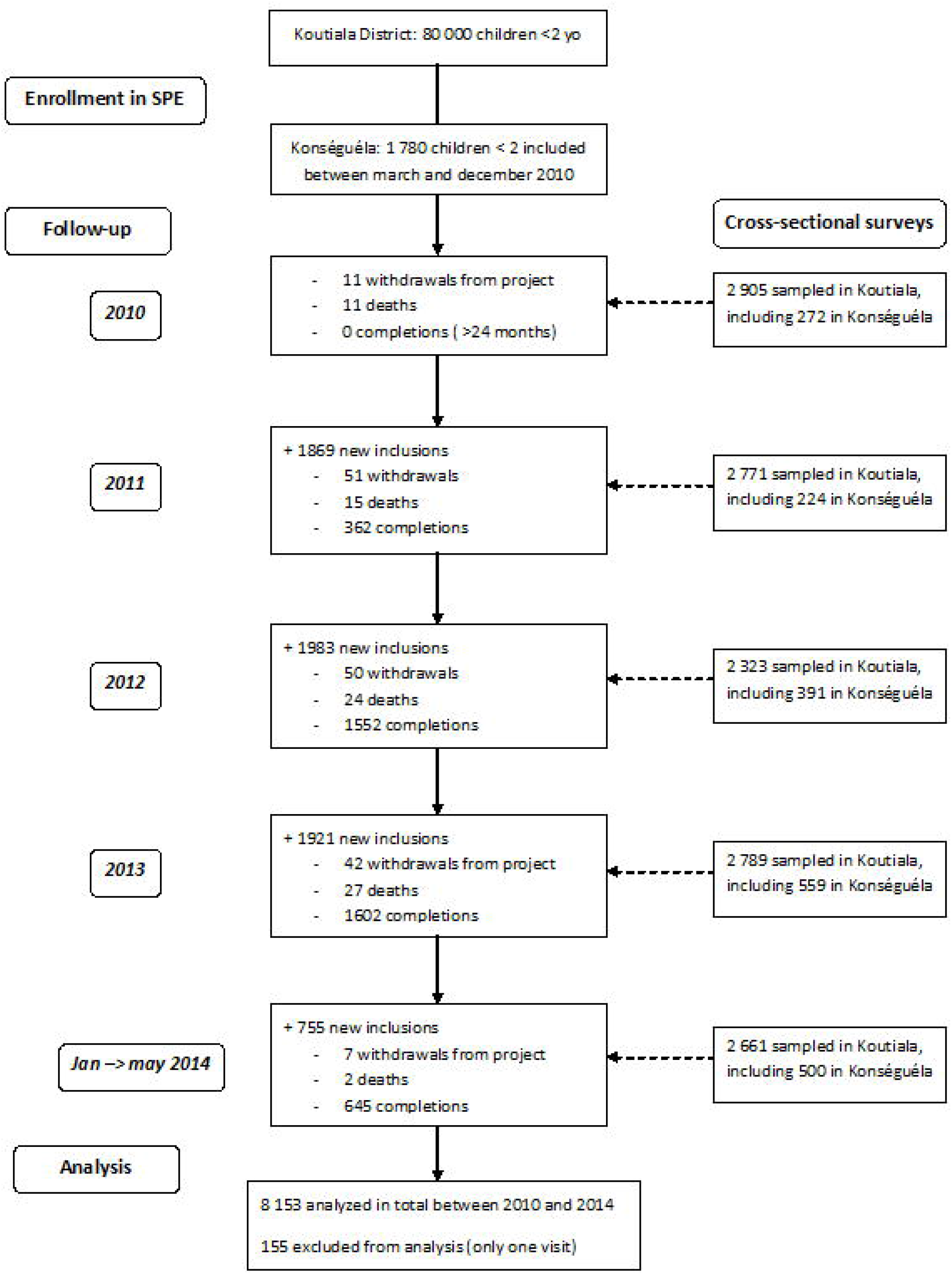
Timeline of the SPE project, Mali, 2010-2014.

For malaria, a network of village-based malaria health workers performed early detection using rapid diagnostic tests, treatment of uncomplicated cases, and referral to CSCOM for complicated cases. These teams are comprised of community members and trained on their tasks by the MoH. MSF additionally provided capacity building and phone credits to ensure communication.

Additionally, Seasonal Malaria Chemoprophylaxis (SMC) was implemented 2012 onwards in the whole district of Koutiala, including Konséguéla, for children 3-59 months of age, using Fansidar-Amodiaquine as an intermittent treatment (one dose every month for three months) following WHO recommendations for areas of highly seasonal transmission.

### Program monitoring

Program monitoring data were obtained from health booklets and recorded into an electronic database by data clerks present on site. Data included sex, age, vaccination status, anthropometric measurements from all visits, Plumpy Doz distribution, bednet distribution, morbidities reported by the mother between visits (diarrhea, malaria, pneumonia), hospitalization history and health status at discharge (death, loss to follow-up or healthy). Data were routinely monitored on site to ensure high quality standards. Community health workers were contacted when a child missed a visit; corresponding reasons for his or her absence or the date of death, if appropriate, were recorded.

Data from the booklets were entered daily by trained data entry clerks, when the mothers were bringing the child to the appointment. A data manager checked for inconsistencies, missing values and outliers on a weekly basis. Twice per month, mother and child registers were cross-checked with the database for quality control.

### Cross-sectional surveys

In April 2010, a district-wide cross-sectional survey was conducted to provide baseline information on children under five.

Additional cross-sectional surveys in the district of Koutiala were conducted March 2011, May 2012, April 2013 and June 2014 (see fig.1). In these surveys, an average of 2 500 children aged 0 to 24 months were surveyed using cluster-based sampling with 150 clusters in about 120 villages selected proportionally to population size[30]. After the baseline survey, the 17 villages of the Konséguéla health area were included into the survey sample and accounted for 300 children on average to allow for comparison between the health area and the rest of the district.

In each cluster, 40 children aged 0 to 59 months were randomly selected following the Expanded Program for Immunization (EPI) random walk method[31].

Informations on age, height, weight, middle-upper-arm circumference, possession of vaccination card and vaccine administration were collected. For nutritional indicators, we compared global and severe acute malnutrition prevalence according to WHO definitions (weight-for-height Z-score<-2 or Middle-Upper-Arm-Circumference (MUAC)<125mm; weight-for-height Z-score<-3, MUAC<115mm or presence of bilateral oedema for SAM). The prevalence of stunting for both populations was also calculated. Evolution of height-for-age Z-scores was analyzed over the course of the child’s growth during their participation in the program.

Coverage of distributed bednets was defined as the number of bednets actually distributed among the children eligible to bednet distribution, either at inclusion or discharge. RUSF coverage was computed as the number of children receiving the full amount of RUSF distributions.

Surveys were carried out during the same time of the year, before the hunger gap, with comparable standard deviations and cluster-effects used as hypothesis for sample size calculation. Since surveys were not powered a-priori for age stratification, yearly data for children 6 to 24 months of age children were pooled. This allowed for a difference-in-difference analysis comparing children in Konséguéla to those living outside that health area.

One of the main purpose of the SPE program is to prevent stunting, which usually strikes children during this critical period of growth (between 6 and 24 months). This preventive aspect should be reflected among the children completing the program when compared to children of the same age in the rest of the district. Thus, children completing the SPE program were pooled (between 2010 and 2014) and compared to children aged 22 to 25 months when surveyed (pooled over surveys) in the rest of the district.

ANOVA F-tests were used to compare health indicators between groups at different points; Cochran-Armitage tests were used to compare trends over time in the surveys, exact Fisher tests were used to compare proportions. Data were entered and monitored with Epidata 3.1® (Odense, Denmark); all analyses were performed using Stata 12.0® for Windows (College Station, Texas, USA).

#### Costs

The global cost of the comprehensive package was evaluated by adding all expenses over a calendar year on RUSF, vaccines, drugs and treatments used in the CSCOM, ITN bednets purchased, and finally including all wages involved. The cost per child was then calculated by dividing the global cost by the global number of beneficiaries followed over the year.

### Ethical Considerations

The project was approved and supported by the Ministry Of Health, through a Memorandum of Understanding. The surveys received approval from the Ethics Committee of Mali.

Detailed information on the project was provided to parents who agreed to participate in the SPE project. Oral informed consent was obtained.

All children within and outside Konséguéla benefited from the SMC strategy through MSF activity and the EPI strategy of the national health system. Children had access to standard curative care provided by MSF through a partnership with the Ministry of Health in the five CSCOMs and the district hospital in Koutiala.

## Results

Cross-sectional surveys were conducted in the district of Koutiala (including Konséguéla) in April 2010, March 2011, May 2012, April 2013 and June 2014. An average of 2 300 households were sampled, for an average of 2 700 children under the age of two years in each survey (global results in Table 1).

**Table 1.** Demographic and morbidity indicators in under 2 children, Koutiala District (incl Konséguéla), Mali, 2010-2014.

| **Children < 2 years** | **April**<br>**2010** | **March**<br>**2011** | **May**<br>**2012** | **April**<br>**2013** | **June**<br>**2014** |
| --- | --- | --- | --- | --- | --- |
| Households Interviewed (n) | 2 088 | 2 403 | 1 940 | 2 532 | 2 538 |
| Children Interviewed (n) | 2 905 | 2 771 | 2 323 | 2 789 | 2 646 |
| Male (%) | 50.1 | 52.7 | 50.9 | 50.6 | 50.5 |
| GAM (%) | 20.9 | 17.2 | 18.2 | 19.1 | 17.2 |
| SAM (%) | 8.2 | 4.9 | 5.6 | 5.3 | 4.8 |
| Stunting (%) | 35.0 | 39.5 | 36.5 | 32.4 | 33.1 |
| Severe Stunting (%) | 14.9 | 13.7 | 12.4 | 11.1 | 12.2 |
| Vaccination Card present (%) | 62.6 | 59.1 | 58.3 | 57.1 | 59.9 |
| Mortality Rate (/10000/day) | 1.29 | 0.57 | 0.52 | 0.56 | 0.24 |

Participation in the SPE project (children reaching 24 months) was 83.8% (362 / 432) in 2011, 96.4% (1550 / 1609) in 2012, 96.1% (1 602 / 1 667) in 2013 and 98.6% (645 / 654) in 2014 (as of june).

### Acute Malnutrition

Results of the difference-in-difference analysis of MUAC, Weight-for-Height Z-score, Height-for-Age Z-score, GAM and SAM prevalences are shown in table 2.

**Table 2.** Difference in difference analysis: results for Acute Malnutrition indicators, Mali 2010-2014.

|  | Period | Koutiala | Konseguela | Difference<br>Kou vs Kon<br>(p-value) | Difference-in-Difference<br>(p-value) |
| --- | --- | --- | --- | --- | --- |
| Muac (mm) | April 2010 | 144.31 | 144.64 | 0.33 (0.611) | 1.21 (0.078) |
|  | 2011-2014 | 145.48 | 147.02 | 1.54 (0.001) |  |
| WfH Zscore | April 2010 | -0.936 | -0.842 | 0.093 (0.476) | 0.08 (0.545) |
|  | 2011-2014 | -0.971 | -0.794 | 0.177 (0.001) |  |
| GAM (%) | April 2010 | 17.39 | 12.80 | 4.59 (<0.001) | -0.49 (0.22) |
|  | 2011-2014 | 15.06 | 10.96 | 4.10 (<0.001) |  |
| SAM (%) | April 2010 | 5.01 | 2.60 | 2.41 (<0.001) | -0.42 (0.17) |
|  | 2011-2014 | 3.57 | 1.58 | 1.99 (<0.001) |  |

All indicators are found to be significantly lower in Konséguéla than in the rest of the district. Nonetheless, for every indicator, the difference-in-difference changes between the baseline and the last survey and between were not significantly different between the two areas.

Noticeably, the coverage of fully distributed RUF was 92% (4 049 / 4 401) for children completing the program.

### Stunting

There was no meaningful change in the prevalence of stunting over time in children followed up for 18 months in Konséguéla and in the rest of the district.

Some differences, however, were noted in the growth of children overall. The HAZ mean for children completing the SPE program (Konséguéla) was −1.65, which corresponds to a prevalence of stunting in the cohort of 35.9% (see table 3). Stunting prevalence in the district without Konséguéla consistently remained above 50%, for an HAZ mean of −1.97 among children 22 to 25 months old (pooled over surveys). This 20% difference in HAZ means and the difference in stunting were statistically significant (p<0.001). Linear growth curves showed the same pattern, as children living outside of Konséguéla had a mean height of 79.3 cm, whereas those in Konséguéla were on average 81.5 cm tall at completion of the program. The ensemble of these health interventions seems to preserve approximately 0.5 Z scores in height-for-age, or 2 cm of growth.

**Table 3.**
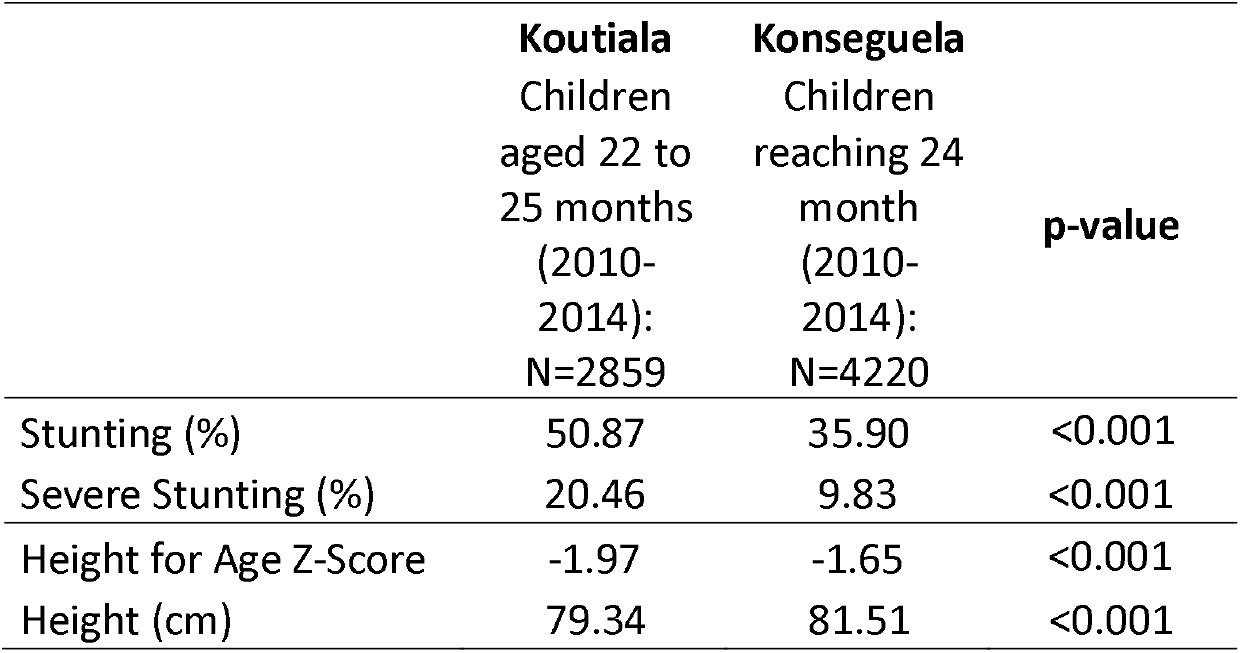
Stunting and growth: comparison of pooled children, Mali, 2010-2014.

### Mortality

Mortality rates computed in the prospective cohort in Konséguéla significantly decreased over time, though changes were not statistically significantly (p=0.06). Mortality rates did not change in the rest of the Koutiala district (p=0.48). The difference-in-difference analysis showed a significantly stronger decrease in the mortality rate in Konséguéla than in Koutiala (p=0.04; see table 4).

**Table 4.** Difference in difference Analysis: Mortality rates, Mali, 2010-2014.

| Period | Koutiala |  | Konséguéla |  | Difference<br>Kou vs Kon<br>(p-value) | DID<br>Change<br>from 1st<br>period<br>(p-value) |
| --- | --- | --- | --- | --- | --- | --- |
|  | † / N | Rate<br>(/10000/day) | † / N | Rate<br>(/10000/day) |  |  |
| 15 Jul 2010-15 Mar 2011 | 20 / 2066 | 0.41 | 13 / 1345 | 0.40 | 0.99 | -0.21 (0.04) |
| 15 Sep 2011-15 May 2012 | 16 / 1574 | 0.33 | 10 / 3367 | 0.11 | <0.001 |  |
| 15 Aug 2012-15 Apr 2013 | 16 / 1719 | 0.34 | 12 / 3656 | 0.12 | <0.001 |  |
| 1 Oct 2013-1 June 2014 | 13 / 1613 | 0.25 | 9 / 3629 | 0.09 | 0.02 |  |

### Vaccination and Malaria

Vaccination coverage for all antigens delivered within the program is described in table 5. The coverage for every antigen increased over time in Konséguéla, while remaining stable in the overall district. The change was significant according to the difference-in-difference analysis (p<0.001).

**Table 5.** Difference in Difference Analysis: Vaccination coverages for main antigens, Mali, 2010-2014.

| Antigen | Pentavalent<br>(3 doses at 24m) |  | Measles<br>(1 dose at 11m) |  | Poliomyelitis<br>(3 doses at 24m) |  |
| --- | --- | --- | --- | --- | --- | --- |
|  | Koutiala | Konséguéla | Koutiala | Konséguéla | Koutiala | Konséguéla |
| Coverage (%) in April 2010 | 44.6 | 36.9 | 37.8 | 25.2 | 43.6 | 39.0 |
| March 2011 | 40.6 | 57.2 | 43.3 | 34.7 | 40.8 | 64.1 |
| May 2012 | 41.2 | 72.1 | 41.4 | 77.4 | 43.1 | 69.6 |
| April 2013 | 39.6 | 61.0 | 34.7 | 61.4 | 43.3 | 65.6 |
| June 2014 | 42.0 | 67.9 | 47.2 | 55.9 | 44.5 | 67.2 |
| Difference 2010-2014 (p-value) | 0.79 | <0.001 | 0.35 | <0.001 | 0.91 | <0.001 |
| DID change from April 2010 (p-value) | 30.9 (0.001) |  | 36.7 (0.001) |  | 28.3 (0.001) |  |

**Table 6.** Distribution of Costs for Comprehensive Pediatric Package, Mali, 2010-2014.

|  | 2010-2014 | 2015-now |
| --- | --- | --- |
|  | Full package with Plumpy<br>Doz (RUSF) 250/kcal/d | Switch RUSF to 120 kcal/d |
| EPI antigens (storage, shipping, human resources) | 27\$ | 27\$ |
| Routine consultations (6 planned, including staff incentives etc.) | 30\$ | 30\$ |
| Sick consultations (average of 6 over 2 years) | 24\$ | 24\$ |
| Bednets (1 at inclusion, 1 at completion) | 7.3\$ | 7.3\$ |
| RUSF (including storage and logistics) | 73\$ | 32,5\$ |
| SMC costs (including human resources, drugs, logistic aspects) | 30\$ | 30\$ |
| **Total cost per child (over 2 years)** | **191.3\$** | **151.8\$** |
| **Cost per child per month** | **8.0\$** | **6.3\$** |

Malaria episodes between visits are not reported due to insufficient data.

Bed net coverage was 99% (8 170 / 8 243) at inclusion between March 2010 and June 2014, and 89% (3 947 / 4 401) at discharge, as of June 2014.

### Costs

The comprehensive pediatric package cost USD 95 per child for a year, exclusive of costs of hospitalization and treatment of acute malnutrition. 56% of this cost was due to the food supplement. Costing of both well-child and sick visits includes salary incentive for personnel, but not the Ministry of Health salary.

## Discussion

In this case study, results of cross-sectional surveys and program monitoring suggest a significant and sustained impact of a community based, comprehensive health package on major child health indicators. Most notably, decreases in mortality were more pronounced in the intervention area. Trends for other indicators suggest additional benefits.

Consistent with sustained efforts to reduce poverty and improve the overall health of children in Mali, infant mortality rates decreased in the district as a whole between 2010 and 2011. Nutritional status may serve as a more sensitive indicator (Pelletier[32-34] and Lutter and al.[35]) of child health in the context of low and decreasing mortality. Here, we see a stabilization in nutritional status with fluctuations between years while mortality decreased. Moreover, children reaching 24 months in Konséguéla were significantly less stunted and taller than their counterparts outside of the health area.

The SPE project was highly accepted by the population, with more than 98% of the children completing the program in 2014. Through greater attendance to health visits, coverage for all EPI antigens improved significantly over the study period, and did so compared to little to no change elsewhere in the district. In Konséguéla, coverage improved by between 64% and 150% for different EPI antigens between 2010 and 2014, whereas the coverage remained relatively stable in the rest of the district (below 45% for each antigen). Other studies have shown that bimonthly visits to villages by outreach teams also have pronounced effects on vaccine coverage (13 to 16% increase in DPT3 coverage was shown by Ryman and al.[27,28]).

Standards of care in the non-intervention area are constantly improving through reinforcement of management of malnutrition and EPI activities in the four CSComs supported by MSF. Similarly, SMC is delivered every year to all children under 3 in the whole Koutiala district. SMC campaigns were followed by coverage and adherence surveys that also provided useful information about malaria episodes and hospitalizations in this population.

Sharing and resale of the RUF product were seldomly observed within the community throughout the study, and appear to occur only in the town of Konséguéla. Spillover effect (ie. residents of villages outside the Konséguéla Health Area trying to access the intervention package) was non-existent, due to the organization of the SPE program: every mother and her child was clearly identified at inclusion and individually followed-up for 18 months. When a mother from another health area would consult to the health center, she could not be included in the program without the individual booklet for her child.

These findings are limited by several factors. Prospective data were only collected on children participating in the SPE program in Konséguéla, and not in the rest of the district. Thus, comparison of indicators between the intervention and the rest of the district were done by cross-sectional surveys, and consequently are limited by this design. Despite being repeated yearly, causality cannot be inferred from survey findings. Furthermore, findings cannot be generalized beyond the targeted area without additional information. Although child mortality was a main objective, the difference found between the two areas could be due to lack of power (the event is so rare that the sample size required to detect statistical differences exceeds the population of the Konséguéla health area). Moreover, data on probable causes of death were lacking while information about malaria episodes was not consistently collected. For ethical and programmatic reasons, additional antigens (i.e. PCV) and interventions (i.e. SMC) were introduced during the 3 year course of the program, resulting in evolving or incomplete data for some objectives, and complicating interpretation of the results. Changes, however, applied to the entire district.

These shortcomings highlight the need for further research with adequate study designs to formally test some of the hypotheses assumed in this case study. For example, a cluster randomized design with villages receiving different paediatric packages would be useful moving forward. Furthermore, cost-effectiveness analysis would also be valuable for policy-makers selecting between different intervention packages.

To replicate and scale up the program to the greater district, which is the following step in the near future, costs need to be considered. The program did change in the course of the 5 years, so only the last 2 years were taken into account here: the package reported here cost around USD 95 per child for one year, of which more than half is the cost of RUSF. Scaling up the intervention would induce an unsustainable rise in those costs, thus other options are required. Several ready-to-use complementary food supplements are currently under development or are already available [36-39]. Switching the RUF to a less expensive product with similar characteristics to the one utilized was the next logical step, as it was finally decided in the course of 2014. If similar health and growth outcomes can be obtained, the cost of the comprehensive package would decrease to approximately USD 75 per child for a year, reducing the total intervention costs by 23% and making scale-up more feasible.

Refrigeration costs for vaccination should also be considered. If flexibility can be added to the cold chain, even at the last stage prior to vaccination, costs associated with equipment, fuel and maintenance of cold chain equipment could be potentially reduced[40,41].

The improvement in the nutrition and vaccination indicators in the intervention area compared to the larger district is most likely a direct result of high coverage. The challenge now is to devise delivery methods at lower cost per child, that maintain coverage levels of ≥ 90% for nutritional supplementation, malaria prevention and treatment as well as EPI on a scale ten times higher than this pilot project.

Human resource challenges can be partially alleviated through utilization of properly trained and paid community health workers networks. Those networks are real pillars of the Malian civil society and are present in every town in the country, which should make implementing programs such as these on a larger scale feasible.

## Data Availability

All study databases are available upon request to the corresponding author.

